# Mathematical model suggests current CAR-macrophage dosage is efficient to low pre-infusion tumour burden but refractory to high tumour burden

**DOI:** 10.1101/2025.08.28.25334697

**Authors:** Shilian Xu, Maoxuan Liu

## Abstract

Chimeric antigen receptor (CAR)-macrophage therapy is a promising approach for tumour treatment due to antigen-specific phagocytosis and tumour clearance. However, the precise impact of tumour burden, dose and dosing regimens on therapeutic outcomes remains poorly understood. We developed ordinary differential equation (ODE) mathematical modelling and utilised parameter inference to analyse *in vitro* FACS-based phagocytosis assay data testing CD19-positive Raji tumour cell against CAR-macrophage, and revealed that phagocytosing efficiency of CAR-macrophage increases but saturates as both Raji cell and CAR-macrophage concentrations increase. This interaction resulted in bistable Raji cell kinetics; specifically, within a particular range of CAR-macrophage concentration, low tumour burdens are effectively inhibited, while high tumour burdens remain refractory. Furthermore, our model predicted that CAR-macrophage dosages typically suggested by current clinical trials yield favourable therapeutic outcomes only when tumour burden is low. For split CAR-macrophage infusion with fixed total dosage, the first infusion with high CAR-macrophage dose delivers superior treatment outcomes. Finally, we identified alternative infusion regimens: five billion cells administered monthly for three months, or seven billion cells every two months for six months, can efficiently suppress Raji cell replication irrespective of tumour burden. Our findings highlight CAR-macrophage therapeutic outcomes are strongly influenced by both tumour burden and different dosing regimens. This work underscores that reducing tumour burden, increasing CAR-macrophage dose in the first infusion and prolonging CAR-macrophage persistence are key strategies for achieving durable responses.

**Highlights (mandatory):**

- Our model demonstrates that ongoing CAR-macrophage dosage is efficient to low tumour burden but refractory to high tumour burden.
- CAR-macrophages are predicted to persist *in vivo* for over 60 days according to our model framework
- CAR-macrophage mediated tumour phagocytosing kinetics increases and saturates as both CAR-macrophage and Raji cell concentrations increase.
- With same infusion cell number, single-dose infusion provides better therapy outcomes than split-dose infusion does for 30-day post-infusion.
- Multi-dose CAR-macrophage infusion regimens achieve superior long-term tumour control compared to single large-dose infusions independent of tumour burdens.

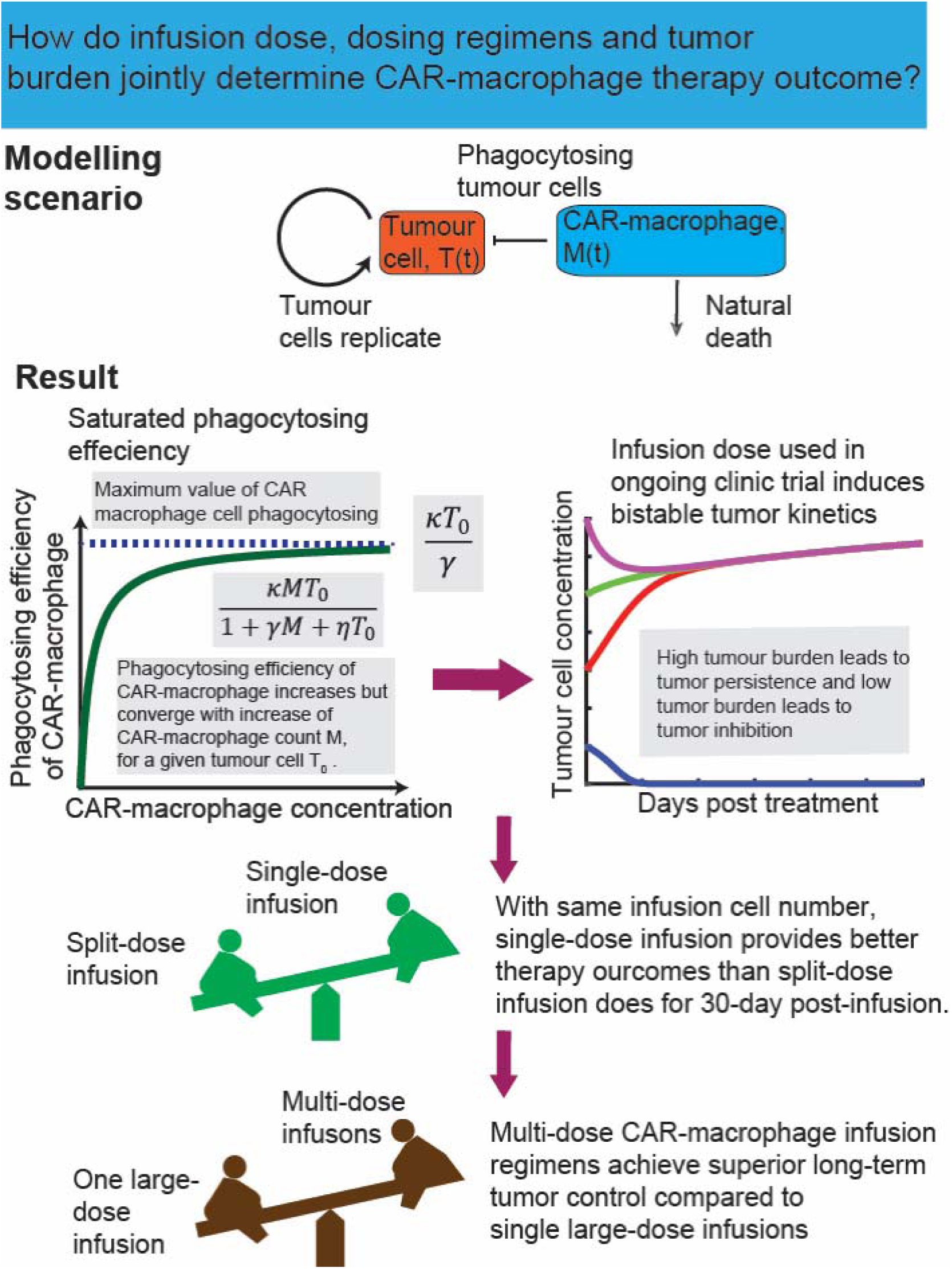
Graphical Abstract (optional)

## Introduction

Adoptive cell therapy, a field that harnesses the patient’s own immune system to fight diseases, has emerged as a promising approach for cancer treatment. A cornerstone of this field is the engineering of chimeric antigen receptors (CARs), which confer new antigen-specific capabilities to T cells [1], natural killer cells [2] and macrophages [3] a new ability to lyse tumour cells or virus infected cells [4, 5].

Chimeric antigen receptor T cells (CAR T-cells) represent a revolutionary advance in cancer treatment, particularly for leukemias and lymphomas, and are often referred to as “living drugs” [6] for patients who failed chemotherapy and radiation therapy.

CAR-T cell therapy has been approved by the FDA (US Food and Drug Administration) to treat B-cell acute lymphoblastic leukemia (B-ALL), diffuse large B cell lymphoma (DLBCL), mantle cell lymphoma (MCL), and multiple myeloma (MM) [7]. It has demonstrated remarkable rates of complete remission, including 76% in B-ALL [8], 54% in DLBCL [9] and 89% in MM [10]. Despite these successes, CAR-T cell therapy is limited by significant challenges, including high costs, the risk of severe side effects such as cytokine release syndrome, and a lack of efficacy against solid tumors due to the immunosuppressive tumor microenvironment [3].

Given these limitations, there has been a growing interest in engineering other immune cells for cancer therapy. Macrophages, as crucial innate immune cells, are particularly attractive candidates due to their intrinsic capacity for phagocytosis, their ability to actively infiltrate tumors, and their role in modulating the adaptive immune response [11]. The development of CAR-macrophages (CAR-M) represents a new frontier in immunotherapy. Although Reinhard Andreesen et al demonstrated the feasibility and safety of infusing high doses of autologous monocyte-derived macrophages, they did not demonstrate antitumor efficacy [12, 13]. Several studies have emerged since 2018, aiming to direct macrophage antitumor function towards tumour cells [14–17]. For instance, Liu et al demonstrated CAR-macrophages and CAR-T cells cooperatively lysed Raji cells (a human CD19-positive Burkitt’s lymphoma tumour cell line) *in vitro* [17]. Lei et al demonstrated CAR-macrophages exhibit superior orthogonal phagocytosis and antitumor functions in treating malignant gliomas [14]. Klichinsky et al. initiated a Phase 1 clinical trial (NCT04660929) for CAR-macrophages in the treatment of HER2-overexpressing solid tumours, which aims to assess objective response (either a complete response (CR) or partial response (PR)) in subjects who received at least one CAR-macrophage infusion [15, 18]. However, the foundational mechanisms governing the efficacy of CAR-macrophage therapy—specifically, how factors like tumor burden, dose and dosing regimens influence outcomes—remain incompletely understood.

Mathematical modelling offers a powerful approach for elucidating the complex dynamics of cancer and immunotherapy [19–22], including monoclonal antibody [23], CAR-T cell [20, 22] and bispecific antibody [24–26], and explained variations in therapeutic outcomes. For example, mathematical models have explained the observed phenomenon that patients with higher tumour burdens at the start of CAR-T treatment are less likely to both attain and maintain a deep response compared to those with a lower tumour burden in B-ALL (unpublished result).

Bispecific antibody induced bistable acute myeloid leukemia (AML) kinetics explains the observed phenomenon that bispecific antibody was less efficacious at high tumour burden even with enough activated cytotoxic CD8+ T cells [26]. Biologically, these phenomena are known as ‘inoculum effect’ [27–29], where high leukocyte counts in patients with B-ALL and AML are strongly negatively correlated with prognostic indicator in terms of remission induction, remission duration, and survival [30]. However, it is unclear whether this phenomenon also applies to CAR-macrophage therapy, and if so, how it might shape therapeutic outcomes.

To understand how tumour burden, CAR-macrophage dosage and dosing regimens jointly determine therapeutic outcomes, we developed a family of two-dimensional mathematical models that involve CAR-macrophage phagocytosis and Raji cell replication. These models were used to analyse *in vitro* FACS-based phagocytosis assay data testing Raji cell against CAR-macrophage, as proposed in Reference [17]. The models allowed phagocytosing efficiency of CAR-macrophage to increase and then saturate as both Raji cell and CAR-macrophage concentrations increase. We revealed that CAR-macrophage and Raji cell interaction leads to bistable Raji cell kinetics, where low tumor burdens are effectively cleared while high burdens are refractory to therapy. Furthermore, our analysis indicates that CAR-macrophage dose, dosing regimens and therapy duration as informed by current clinical trials (CAR-macrophages for the Treatment of HER2 Overexpressing Solid Tumours [18]) can yield favourable therapeutic outcomes only when pre-infusion tumour burden is low. Finally, we identified alternative and more robust dosing regimens: five billion cell infusion administered monthly for three months, or seven billion cell infusion every two months for six months, can efficiently suppress Raji cell replication irrespective of tumour burdens. We also found that while normal macrophages can induce bistable Raji cell kinetics, their tumour-phagocytosing capability cannot suppress Raji cell replication within typical physiological macrophage concentration ranges. These results revealed that CAR-macrophage therapeutic outcomes are strongly influenced by tumour burden, doses and dosing regimens. Furthermore, reducing tumour burden and prolonging CAR-macrophage treatment duration yields improved therapeutic outcomes. This study has significant implications for the development and optimization of CAR-macrophage therapies.

## Result

### Model Overview: Monostable vs Bistable Tumour Kinetics

We investigated two types of tumour kinetics-monostable and bistable tumour kinetics-to describe the relationship among tumour cell concentration, CAR-macrophage concentration, and tumour burden. **Monostable** tumour kinetics refers to a scenario where tumour persistence or eradication is solely determined by the CAR-macrophage concentration, irrespective of tumour burdens (Fig.1A).

**Figure 1.**
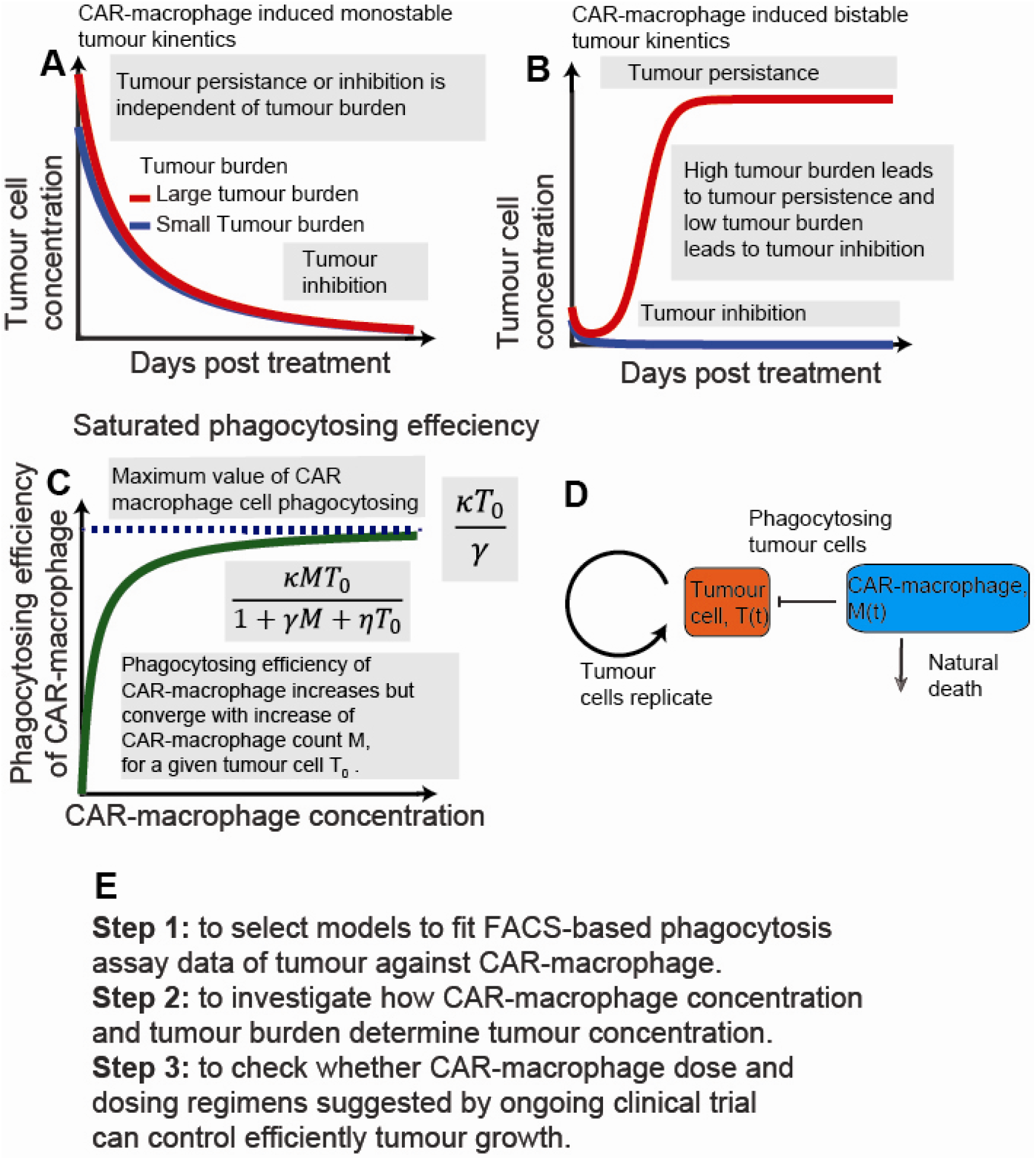
**Model Overview: Illustrating Tumour Kinetics, Saturated CAR-Macrophage phagocytosing Efficiency, and Model Structure. (A) Monostable tumour kinetics**: In the scenario, persistence or eradication of tumour is solely determined by the CAR-macrophage concentration, irrespective of tumour burdens. **(B) Bistable tumour kinetics**: Within a specific CAR-macrophage concentration range, tumour kinetics with high tumour burdens persist, while those with low tumour burdens are suppressed. **(C) Saturated CAR-macrophage phagocytosing efficiency**: phagocytosing efficiency of CAR-macrophage increases but saturates with both tumour cell and CAR-macrophage concentration. **(D) Model schematic diagram**: tumour kinetics in the presence of CAR-macrophages is divided into two compartments, including tumour replication kinetics and tumour loss due to CAR-macrophage phagocytosis. This model does not account for CAR-macrophage proliferation in patients. **(E) Model workflow**: a visual representation of the overall model process.

Alternatively, **bistable** tumour kinetics exhibits tumour persistence and eradication coexisting within the same CAR-macrophage concentration range. Specifically, within a given CAR-macrophage concentration range, tumor kinetics with low tumour burdens are suppressed, while those with high tumour burdens persist (Fig. 1B). In addition, our model reveals that the phagocytic efficiency of CAR-macrophages increases with both tumor cell and CAR-macrophage concentration, but eventually saturates (Fig. 1C). We hypothesise that this saturation in CAR-macrophage phagocytosis can trigger CAR-macrophage induced bistable tumour kinetics between persistence and eradication. It is important to note that unlike CAR-T cells, which undergo massive clonal proliferation when simulated with tumour cells/ target cells [31–33], Unlike CAR-T cells, which undergo massive clinal proliferation, CAR-macrophages do not proliferate *in vitro* or *in vivo* [34–36]. Patients can only receive a limited number of CAR-macrophage therapies, which might affect their efficacy [36]. Therefore, we do not consider that CAR-macrophages proliferate in this manuscript (shown in Fig.1D). The model schematic diagram and model workflow are shown in Fig.1D and Fig.1E.

### Raji Cell Replication Kinetics Follows Logistic Growth in the Absence of CAR-Macrophage or Normal Macrophage

FACS-based phagocytosis assays typically measure the phagocytosing efficiency of CAR-macrophage during a short period (one or two days) [17]. However, in *in vivo* treatment, the time scale often spans more than months or even years [37, 38]. To accurately account for this longer timescale, we confirmed that Raji cell replication kinetics, in the absence of CAR-macrophage or normal macrophage, follows logistic growth [22] (delineated by Model 1). We achieved this by fitting the model to Raji cell proliferation assay data [39] (Fig. S1, Section 1.1, *Supplementary material*).

### CAR-Macrophage Mediated Raji Cell Phagocytosing Kinetics Increases and Saturates with Increasing Concentrations of Both CAR-macrophages and Raji Cells

To quantify CAR-macrophage-mediated Raji cell phagocytosis, we performed FACS-based phagocytosis assays testing human CD19-positive tumour cell (Raji cell) against CAR-macrophages. These assays allowed the estimation of Raji cell concentration before and after co-culturing Raji cells and CAR-macrophages (see *Methods*, Reference [17]). Raji cells were incubated with CAR-macrophages or normal macrophages at different effector (E) to target (T) cell ratio (E: T ratios: 2, 1 or 0.5) for 24 and 48 hours (1 and 2 days) [17]. Normal macrophage-mediated phagocytosis kinetics was measured using same method.

First, using Raji cells and CAR-macrophages as an example, we selected a mathematical model to delineate CAR-macrophage mediated phagocytosis kinetics. To establish a quantitative relationship between viable Raji cell concentration and CAR-macrophage concentration, we tested four mathematical models of CAR-macrophage phagocytosis: an unsaturated model (Model 2, *Methods*, modelled by law of mass action, purple curve in Fig.2A and 2B), semi-saturated model with increase of tumour cell concentration (Model 3, *Methods,* modelled by Hill function, green curve in Fig.2A and 2B), semi-saturated model with increase of CAR macrophage concentration (Model 4, *Methods,* modelled by Hill function, light blue curve in Fig.2A and 2B), and saturated model with increase of both tumour and CAR-macrophage concentration (Model 5, *Methods,* modelled by Beddington-DeAngelis functional response, blue curve in Fig.2A and 2B) to describe the rate of change of tumour cell and CAR-macrophage concentrations. Unexpectedly, only saturated model (Model 5) can fit FACS-based phagocytosis assays data (shown blue curve in Fig. 2A and 2B, Table S4), and unsaturated and semi-saturated model cannot (shown in Table S1, S2, and S3). Normal macrophage mediated phagocytosis kinetics summarised in Table S5.

**Figure 2.**
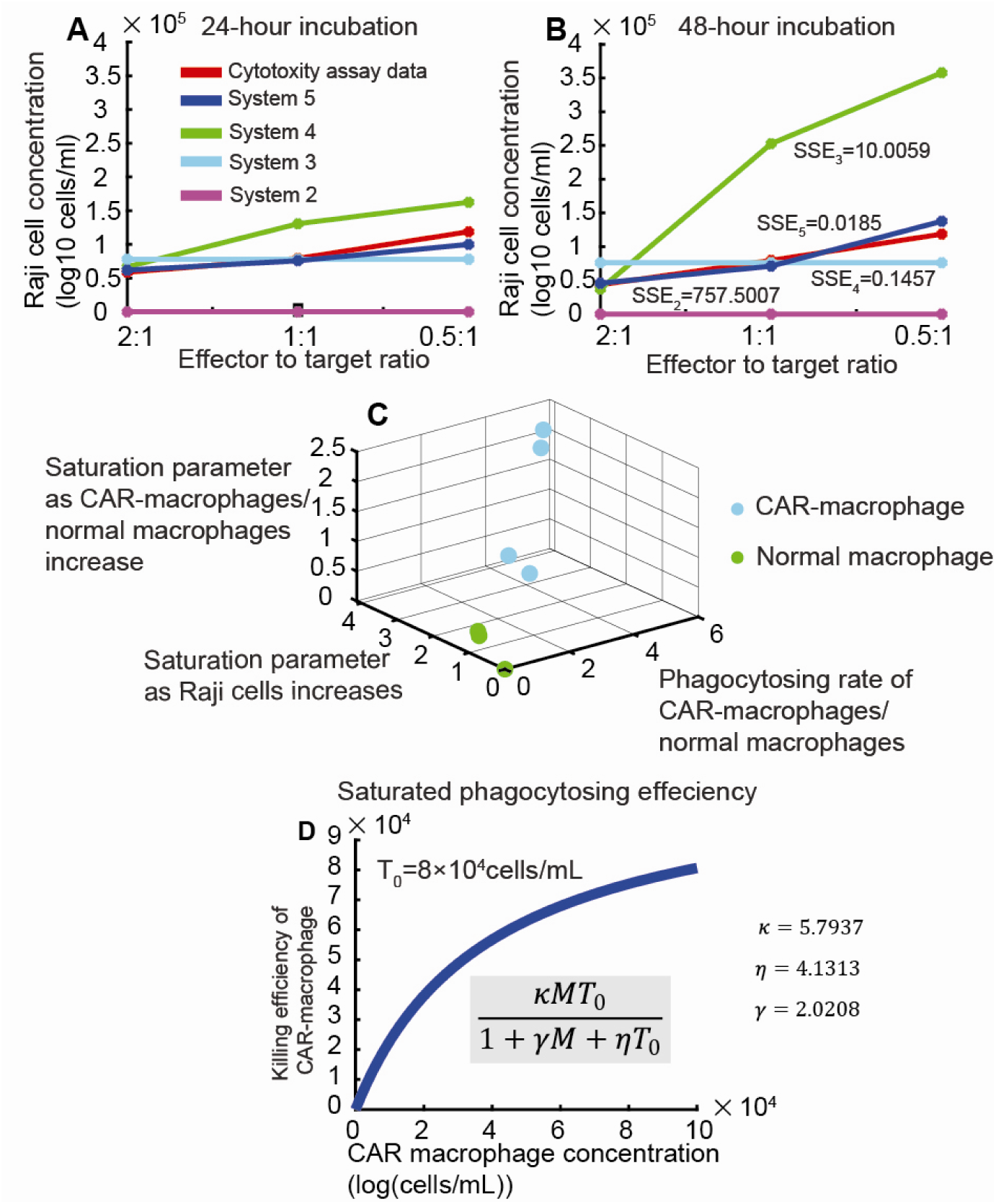
Comparison of FACS-based Phagocytosis Assay Data and Estimated Values, Comparison of Phagocytosing Parameters, and Saturated Phagocytosing Efficiency. (**A** and **B**) **Model Fitting to FACS-Based Phagocytosis Assay Data.** Fitting FACS-based phagocytosis assay data indicates that remaining viable Raji cell concentration increases as the effector-to-target (E: T) ratio decreases (2:1, 1:1, or 0.5:1) at **(A)** 24 hours or **(B)** 48 hours. Purple, green, light blue, blue, and red lines represent Model 2, Model 3, Model 4, Model 5, and the FACS-based phagocytosis assays data, respectively. The Sum of Squared Errors (SSE) for Model 2 (SSE_2_) was 727.5007, for Model 3 (SSE_3_) was 10.0059, for Model 4 (SSE_4_) was 0.1457, and for Model 5 (SSE_5_) was 0.0185. **(C) Comparison of three phagocytosing parameters for CAR-macrophages and normal macrophages.** This panel compares three key phagocytic parameters for CAR-macrophages and normal rate of CAR macrophage or normal macrophage). The y-axis represents macrophage or normal macrophages, highlighting their distinct behaviours. The x-axis represents κ (phagocytosing rate of CAR macrophage or normal macrophage). The y-axis represents *η* (saturation parameter controlling phagocytosing efficiency as tumour cell concentration increases). The z-axis represents γ (saturation parameter controlling phagocytosing efficiency as CAR-macrophage concentration increases). Cyan circles denote phagocytosing parameters for CAR-macrophages, while green circles represent those for normal macrophages. **(D) Simulated Phagocytosis Efficiency.** Phagocytosing efficiency of CAR-macrophages is simulated with estimated parameters. The blue curve represents the saturated phagocytosing efficiency. Parameters used are p = 0.90, K = 7.31 x 10^6^, K= 5.79, 17 = 2.02 and y= 4.13 (the third column in Table S4).

To investigate whether phagocytosing Raji cells leads to additional CAR-macrophage death, we fitted a saturated model that included an additional death rate (Model 6, *Methods*) to fit the FACS-based phagocytosis assays data of Raji cell testing against CAR-macrophages. Surprisingly, we found that the additional CAR-macrophage death rate due to phagocytosing Raji cells is fitted to be zero among all biological replicates (shown in Table. S6), which led us to reject the hypothesis that phagocytosis causes additional CAR-macrophage death. Finally, a comparison of the fitted phagocytosis rates revealed a dramatic difference: phagocytosing rate of CAR-macrophage κ predominantly range from 1 to 6 (Fig. 2C, cyan circle), while that of normal macrophage κ was typically less than 1 (Fig. 2C, green circle). This indicates that the chimeric antigen receptor (CAR) dramatically enhances phagocytic capability of CAR-macrophages. The phagocytosing efficiency of CAR-macrophage is given in Fig. 2D.

### Both CAR-Macrophage and Normal Macrophage Induce Bistable Raji Cell Kinetics

Using bifurcation analysis and numerical simulation of Raji cell kinetics governed by saturated phagocytosing efficiency (Model 5), we found that both CAR-macrophages and normal macrophages induced bistable Raji cell kinetics. **Bistability** means that for a given CAR-macrophage or normal macrophage concentration interval, Raji cell kinetics with small tumour burden are inhibited, while those with large tumour burden persist. In contrast, **monostability** means that the persistence or eradication of Raji cell is solely determined by magnitude of CAR-macrophage concentration, independent of tumour burdens. Furthermore, we refined CAR-macrophage or normal macrophage concentration intervals provided in existed reference.

We observed that CAR-macrophages exhibited a smaller CAR-macrophage concentration interval over which bistable Raji cell kinetics occurred compared to normal macrophages. This finding supports that CAR-macrophages provide superior anti-tumour capability on Raji cell kinetics. Furthermore, we refined CAR-macrophages or normal macrophage concentration intervals provided in existed reference. For example, Reinhard al et used 1.5 x10^9^ cells per infusion [12], and an ongoing clinic trial (CAR-macrophages for the Treatment of HER2 Overexpressing Solid Tumors) used a total of five billion cells [18]. Given that a typical adult has a blood volume of approximately 5 litres, CAR-macrophage concentrations are converted cells/ml by dividing the infusion dosage by 5000 ml.

First, we use the CAR-macrophages as an example. In simulations of Raji cell kinetics with saturated phagocytosing efficiency, a threshold C*=1.4×10^6^ cells/ml existed that divided CAR-macrophage concentration interval into two regimes, shown as a bifurcation diagram (shown in Fig. 3D). For this figure, we used the CAR-macrophage phagocytosing parameters in Column 3 of Table S4, *Supplementary Information*. At different CAR-macrophage concentration intervals, Raji cell kinetics exhibited different dynamics. Specifically, Raji cell kinetics exhibited bistable behaviours within the CAR-macrophage concentration interval between 0 and C*, where Raji cell kinetics with low tumour burdens were inhibited, and those with high tumour burdens persisted at the same CAR-macrophage concentration (shown in Fig.3). The Raji cell concentration threshold (above which the Raji cell concentration is not affected) increased with increase of CAR-macrophage concentration (the red dashed curve in Fig. 3D). For example, at low CAR-macrophage concentration C=3 ×10^5^ cells/ml (Reinhard al et, [12, 13]), tumour burden 1×10^6^ cells/ml, 3×10^6^ cells /ml, 5×10^6^ cells/ml and 7×10^6^ cells/ml all persisted (shown in Fig. 3A and D). In contrast, at high CAR-macrophage concentration 10×10^5^ cells/ml, representing current clinic trial [18], only high tumour burden 3×10^6^ cells/ml, 5×10^6^ cells /ml and 7×10^6^ cells /ml persisted (shown in Fig. 3B and D). At CAR-macrophage concentration higher than the threshold C*, Raji cell was inhibited independent of high tumour burden (shown in Fig. 3C and D). CAR-macrophage kinetics with different dosages were shown in Fig. S2.

**Figure 3.**
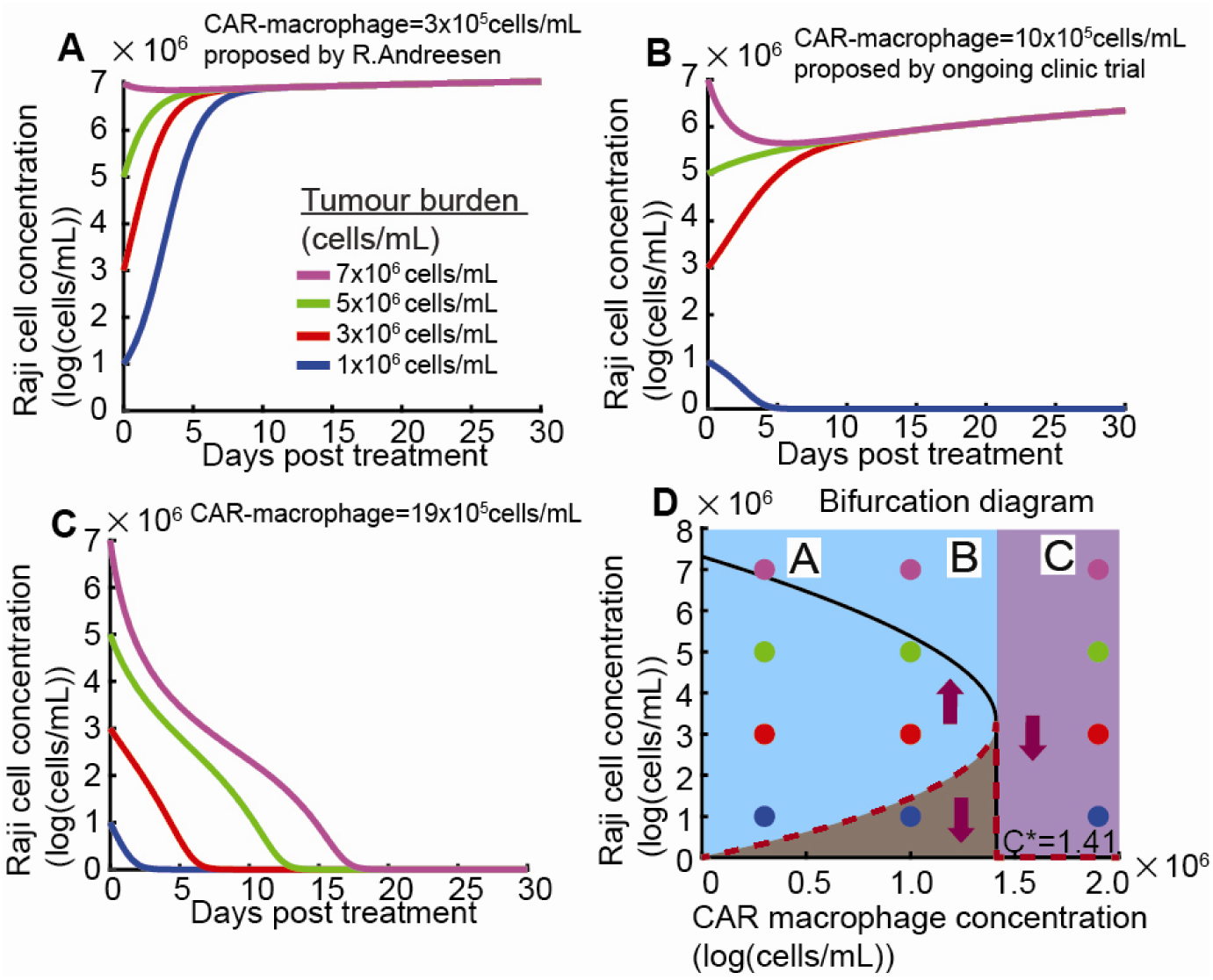
**Simulated Raji Cell Kinetics under Varying Tumour Burden and CAR-Macrophage Concentrations**. **(A-C). Simulated Raji Cell Kinetics.** Raji cell kinetics are simulated at different CAR-macrophage concentrations and tumor burdens, illustrating the concept of **bistability** and **monostability**. **(A)** and **(B)** when CAR-macrophage concentration was between 0 and C*, Raji cell kinetics persists if the tumour burden was above the dashed red curve and were eradicated if the tumour burden was below the dashed red curve. Panels (A) and (B) show specific examples of these dynamics. **(C)** Raji cell kinetics was eradicated independent of tumour burden when CAR-macrophage concentration was greater than C*. **(D) Bifurcation diagram** showing Raji concentration as a function of CAR macrophage concentration. Tumour burden threshold increases with increase of CAR-macrophage concentration (red dashed curve). The maximal Raji cell carrying capacity (black solid curve) decreases with increases of CAR macrophage concentration. Purple arrows illustrate any the trajectory from different tumour burdens. Purple, green, red and blue lines and circles represent Raji cell and CAR-macrophage kinetics with tumour burdens 1×10^6^ cells/ml, 3×10^6^ cells /ml, 5×10^6^ cells/ml and 7×10^6^ cells/ml, respectively, as shown in (A–D).

Similarly, normal macrophages also induce bistable Raji cell kinetics (shown in Fig. 4A). However, the threshold of macrophage concentration for bistability is approximately located at C*=2.68×10^6^ cells/ml, which is approximately 3-8 times higher than that monocyte level in human (2 to 8×10^5^ cells/ml). This high threshold concentration explains why normal macrophages alone are unable to fully suppress Raji cell replication, but can reduce or suppress the tumor burden, a phenomenon observed in previous studies [12, 13] (shown in Fig. 4B).

**Figure 4.**
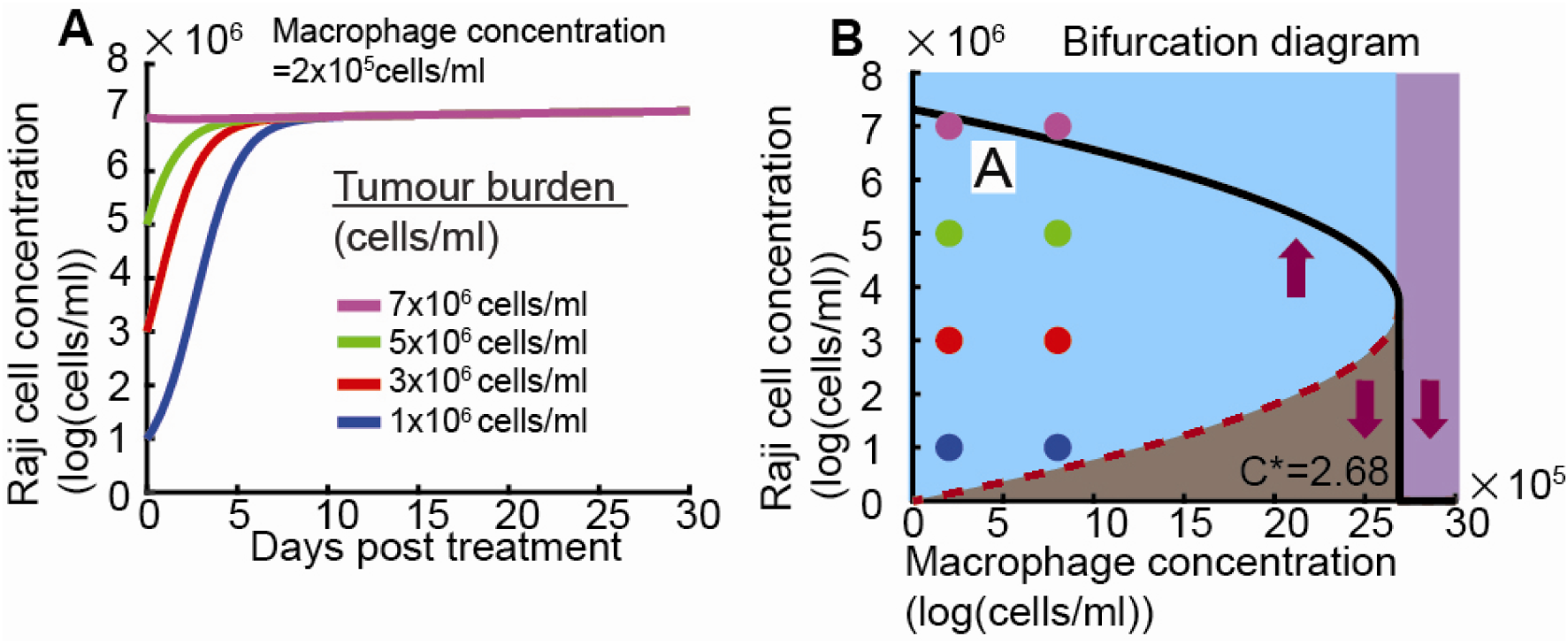
Simulated Raji Cell Kinetics under Varying Tumour Burden and Normal Macrophage Concentrations. (A) Simulated Raji Cell Kinetics. When normal macrophage concentration was between 0 and C*, Raji cell kinetics persisted if tumour burdens were above the dashed red curve and were eradicated if tumour burdens were below the dashed red curve. **(B) Bifurcation diagram** showing Raji cell concentration as a function of normal macrophage concentration. The tumour burden threshold increases with increase of normal macrophage concentration (red dashed curve). The maximal Raji cell carrying capacity (black solid curve) decreases with increases of normal macrophage concentration. Purple arrows illustrate the trajectory from different tumour burdens. Purple, green, red and blue lines and circles represent simulated Raji cell and CAR-macrophage kinetics with tumour burden 1×10^6^ cells/ml, 3×10^6^ cells/ml, 5×10^6^ cells/ml and 7×10^6^ cells/ml, respectively, as shown in (A–B).

### Impact of CAR-Macrophage Dose and Dosing Regimens on Raji Cell Kinetics

Building on our investigation into how CAR-macrophage concentrations affect Raji cell kinetics, this section explores how different clinical dosing regimens determine therapy outcomes. While a single infusion to achieve a high concentration like 19 x 10^5^ cells/ml may not be easily feasible, maintaining high concentrations through multiple infusions presents a promising alternative. We modeled five distinct therapy scenarios to evaluate their impact.

We examined five treatment scenarios, including three derived from the ongoing clinical trial (CAR-macrophages for the Treatment of HER2 Overexpressing Solid Tumors [18]), which categorise single-dose and split-dose infusions:

- **Scenario A** (single-dose infusion, Treatment Scenario One proposed by Reference [18], shown in Fig.5A): a full intravenous dose of up to 5 billion cells total on Day 1 (equivalent to 10 x 10^5^ cells/ml, assuming 5 L blood volume).
- **Scenario B** (split-dose infusion, Treatment Scenario Two proposed by Reference [18], shown in Fig.5A): CAR-macrophages are split into three infusions: up to 500 million cells (equivalent to 1 x 10^5^ cells/ml) on Day 1, up to 1.5 billion cells (equivalent to 3 x 10^5^ cells/ml) on Day 3, and up to 3.0 billion cells (equivalent to 6 x 10^5^ cells/ml) on Day 5.
- **Scenario C** (reverse split-dose infusion, shown in Fig.5A): This scenario reverses the order of Scenario B. Infusion of 3.0 billion cells (equivalent to 6 x 10^5^ cells/ml) on Day 1, up to 1.5 billion cells (equivalent to 3 x 10^5^ cells/ml) on Day 3, and up to 500 million cells (equivalent to 1 x 10^5^ cells/ml) on Day 5.

**Figure 5.**
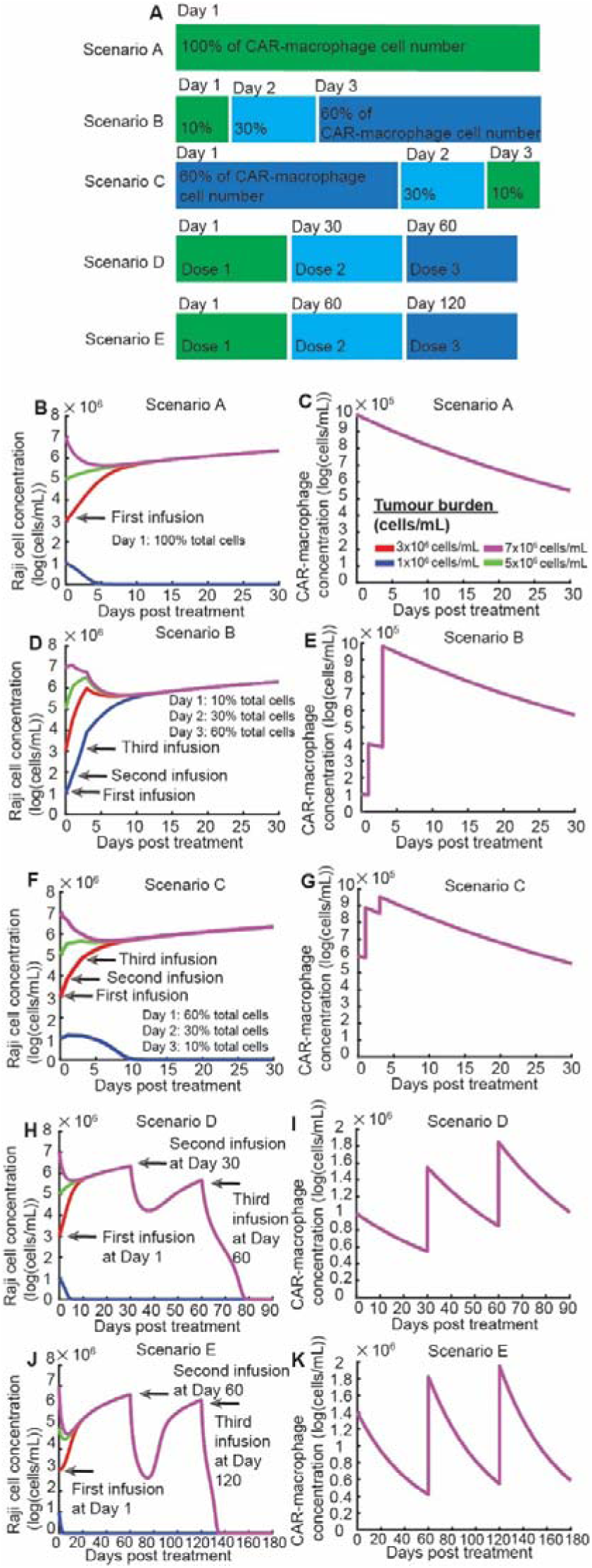
Simulated Raji Cell Kinetics under Various CAR-Macrophage Dosing Regimens. (A) Schematic of Dosing Regimens. This panel illustrates the five simulated therapy scenarios. **Scenario A** provides 100% of the total infused dose (5 billion cells, equivalent to 10 x 10^5^ cells/ml) on Day 1. **Scenario B** provides a split dose (totalling 5 billion cells): 10% on Day 1, 30% on Day 3, and 60% on Day 5. **Scenario C** provides a split dose (totalling 5 billion cells): 60% on Day 1, 30% on Day 3, and 10% on Day 5. **Scenario D** provides single-dose infusions (each at 5 billion total cells, equivalent to 10 x 10^5^ cells/ml) on Day 1, Day 30, and Day 60. **Scenario E** provides single-dose infusions (each at 7 billion total cells, equivalent to 14 × 10^5^ cells/ml) on Day 1, Day 60, and Day 120. **(B-K) Raji Cell and CAR-macrophage kinetics. (B)** and **(C)** Raji cell kinetics and CAR-macrophage kinetics under Scenario A (Single-Dose Infusion). Scenario A, reflecting an ongoing clinical trial regimen, efficiently suppresses Raji cell growth with low initial tumour burden but is not effective against high initial tumour burden. **(D)** and **(E)** Raji cell kinetics and CAR-macrophage kinetics under Scenario B (Split-Dose Infusion). Scenario B in ongoing clinical trial cannot suppresses Raji cell growth independent of tumour burden. **(F)** and **(G)** Raji cell and CAR-macrophage concentration kinetics under Scenario C (Split-Dose Infusion, Reverse Order). Scenario C efficiently suppresses Raji cell growth with low tumour burden but not high tumour burden. **(H)** and **(I)** Raji cell and CAR-macrophage concentration kinetics under Scenario D (Multi-Dose Infusion). Scenario D efficiently suppresses Raji cell growth independent of tumour burden. **(J)** and **(K)** Raji cell and CAR-macrophage concentration kinetics under Scenario E (Extended Multi-Dose Infusion). Scenario E efficiently suppresses Raji cell growth independent of tumour burden. Purple, green, red, and blue lines represent simulated Raji cell kinetics originating from tumour burdens of 1×10^6^ cells/ml, 3×10^6^ cells/ml, 5×10^6^ cells/ml and 7×10^6^ cells/ml, respectively, as shown in (B–K). In Panels (C), (E), (G), (I), and (K), the purple, green, red, and blue curves representing Raji cell kinetics often overlap, reflecting that their CAR-macrophage concentrations are the same across different initial tumour burdens for these specific panels. Parameters used are p = 0.90, K= 7.31 x 10^6^, K = 5.79, 17 = 2.02 and y = 4.13 (the third column in Table S4).

To explore the effects of sustained, high concentrations, we also proposed two additional scenarios.

- **Scenario D (Monthly infusions)**: A total of five billion cells (equivalent to 10 x 10^5^ cells/ml) are infused monthly for three months (shown in Fig.5A).
- **Scenario E (Bimonthly infusion)**: A total of seven billion cells (equivalent to 14 x 10^5^ cells/ml) are infused every two months for a total of six months (shown in Fig.5A).

Then, our simulations reveal a clear correlation among dose, dosing regimens and tumor burden on therapeutic success.

- **Scenario A and C:** Both the single high-dose infusion (Scenario A) and the split-dose regimen with a high initial dose (Scenario C) led to **bistable Raji cell kinetics.** In these cases, CAR-macrophages successfully controlled low tumor burdens but were ineffective against high tumor burdens (Fig. 5B, 5C, 5F, and 5G).
- **Scenario B:** Unexpectedly, the split-dose regimen with a lower initial dose failed to inhibit Raji cell growth, even though the total number of infused cells was the same as in Scenarios A and C (10 x 10^5^ cells/ml, shown in Fig. 5D and 5E).
- **Scenarios D and E**: The sustained, high concentrations achieved in these regimens inhibited Raji cell kinetics regardless of the initial tumor burden (Fig. 5H and 5J). This superior efficacy is directly attributed to the high accumulated CAR-macrophage concentration (Fig. 5I and 5K).

This collective analysis suggests that the infusion dose and dosing regimen, particularly the magnitude of the initial dose, play a decisive role in therapy success. Our model indicates that a higher initial infusion dose provides better outcomes, even when the total infused CAR-macrophage number is consistent.

### CAR-macrophage Persistence

The lifespan of monocytes is relatively short, with a mean of approximately one day [40], whereas that of macrophages is typically long, lasting for months or years [41]. Our simulations shown that after 30 days, the CAR-macrophage concentration remains approximately 0.6×10^6^ cells/ml (equivalent to three billion cells in human, shown in Fig.5B, 5D and 5F). After 60 days, the CAR-macrophage concentration is approximately 0.3×10^6^ cells/ml (equivalent to one and half billion cells in human, shown in Fig.S3A and S3B). This persistence aligns with clinical observations that CAR-macrophages can remain in the host for more than one month in vitro and at least 62 days *in vivo [37, 38]*.

## Discussion

Our analysis, integrating FACS-based phagocytosis assay data with a mathematical model, has yielded several key findings regarding CAR-macrophage therapy. We first demonstrated that the phagocytic efficiency of CAR-macrophages increases and saturates with rising concentrations of both Raji cells and CAR-macrophages. This saturation leads to bistable Raji cell kinetics, a critical finding where therapeutic outcome depends on the initial tumor burden. Specifically, our model predicts that a single high-dose infusion, as well as a split-dose regimen with a high initial dose, can efficiently control low tumor burdens but is refractory to high tumor burdens. In contrast, multi-dose regimens, such as those administered monthly or bimonthly, can inhibit tumor replication regardless of the initial tumor burden due to the high, sustained CAR-macrophage concentrations they achieve. These results highlight that therapy outcomes are not only influenced by tumor burdens but are decisively shaped by the CAR-macrophage **infusion dose** and **dosing regimen**, with a higher initial dose providing better outcomes, even when the total number of infused cells is the same.

The existence of CAR-macrophage induced bistable Raji kinetics has significant implications, especially for clinical trial scenarios such as A and B [18]. From a clinical perspective, these findings underscore the necessity of accurately determining a patient’s tumor burden before initiating CAR-macrophage therapy. When the tumor burden is high, complementary treatments like chemotherapy or radiotherapy may be essential to reduce the tumor burden below the threshold identified in our bifurcation diagram (Fig. 3D). This would allow CAR-macrophage therapy to be effective. As an alternative, our model suggests that higher CAR-macrophage dosages, increased infusion frequency, and a prolonged therapy duration offer a more robust strategy to mitigate therapeutic uncertainty and achieve control independent of tumor burdens. The central finding of our work is that CAR-macrophage concentration can be divided into two intervals: one where tumor burden dictates Raji cell kinetics and another where it does not. Clinically, the ideal approach is to maintain CAR-macrophage concentrations within this second interval, thereby achieving tumor control regardless of burden.

Due to these similarities, interaction between tumour cells and CAR-T cells may also exhibit lysing efficiency of CAR-T cell increases and gradually converges with increasing concentration of either tumour cells or CAR-T cells, thereby inducing bistable tumour cell kinetics. This provides a new mechanism for observed phenomena that patients with higher tumour burdens at the start of CAR-T cell therapy are less likely to both attain and maintain a deep response compared to those with a lower tumour burden in all malignancies, including B cell lymphomas [42].

Unlike CAR-T cells, which undergo massive clonal proliferation when simulated with tumour cells [31–33], CAR-macrophages do not proliferate *in vitro* or *in vivo* [34–36]. In contrast, CAR-T cell proliferation can significantly influence therapy outcomes, potentially overriding the effects of infusion dose and regimen. Because CAR-macrophages do not proliferate, their therapeutic success is almost exclusively dependent on the administered dose and regimen. Our simulations confirm that for single-dose infusions, a higher initial dose provides superior outcomes.

Reinhard Andreesen al et used 1.5 x10^9^ cells per infusion (equivalent to 0.3 x10^6^ cells/ml, assuming 5L blood volume) and found that one patient with more than 25% reduced tumour size and eight patients with 25% increased tumour size [12, 13]. This aligns with our mathematical model’s predictions (Fig. 4A and B); increasing normal macrophage concentration alone cannot suppress tumour replication, but may decrease tumour cell concentration or tumour size. Scenario A and Scenario B, used in current clinical trial, were proposed to suppress HER2 Overexpressing solid tumour growth (for example, breast cancer and stomach neoplasms) [18], and Raji cells are a human B-lineage acute lymphoblastic leukemia (known as a non-solid tumour). First, when comparing the CAR-macrophage dose, dosing regimens, and therapy duration used in Scenario A and Scenario B of the current clinical trial, these parameters may be too low to effectively suppress Raji cell replication. This warrants future research. Next, because macrophages can infiltrate solid tumour tissue and interact with components in tumour microenvironment, it is worthwhile to investigate the role of tumour burden on CAR-macrophage therapy prognosis in HER2-overexpressing solid tumours.

The FACS-based phagocytosis assay is designed to quantify remaining viable tumour cells after incubation with CAR-macrophages and normal macrophages during incubation [17]. Tumour cell and CAR-macrophage mixtures are typically incubated for 24 hours or 48 hours. However, CAR-macrophages can persist in host at least 1 month *in vitro* and at least 62 days *in vivo [37, 38]*. Therefore, results provided by short incubation assays may not reflect the full picture of CAR-macrophage phagocytosing kinetics, as CAR-macrophage dose, treatment duration and dosing regimens determine therapy outcomes. Furthermore, the FACS-based phagocytosis assay only measures tumour cell viability, rather than CAR-macrophage or macrophage cell viability. Our current model suggested that phagocytosing Raji cells does not lead to additional CAR-macrophage death, but this hypothesis requires experimental validation.

Our model, which integrates Raji cell growth parameters from one study and CAR-macrophage phagocytosis parameters from another, successfully predicts the existence of CAR-macrophage-induced bistable kinetics. However, this prediction warrants experimental validation within a single and unified experimental system.

Future work is required to determine the specific bistable concentration interval for a given chimeric antigen receptor (CAR). Additionally, it would be worthwhile to investigate how different CAR designs, combined with monoclonal antibody therapies like Pembrolizumab, influence optimal macrophage dose and dosing regimens. Our current model also assumes a homogenous tumor population; therefore, future modeling should incorporate natural tumor heterogeneity with a mixture of genotypes and varying responses to CAR-macrophage therapy. Finally, because B-lineage acute lymphoblastic leukemia (B-ALL) is known to create a supportive bone marrow microenvironment that promotes tumor survival and resistance, future modeling efforts are warranted to investigate how the tumor microenvironment (TEM) influences CAR-macrophage therapeutic outcomes. This work will further advance our quantitative understanding of how macrophages and combined immunotherapies impact tumor replication.

Immunotherapies aim to kill or phagocytose tumour cells using macrophage, for example, CAR-macrophage and monoclonal antibody (mAb). Anti-CD47 blockade with monoclonal antibody opsonization of cancer cells drive macrophage phagocytosis [43]. For bispecific antibody against acute myeloid leukemia (AML), maintaining high specific antibody concentration and preventing T cell exhaustion are equivalently important to promote and sustain long-term AML control [26]. In the future work, it is worthwhile to quantitively understand how macrophages and monoclonal antibodies determine tumour replication.

## Method

### Mathematical model

To quantitatively describe the relationship between Raji cell concentration and day post-inoculation, we used a logistic growth model (Model 1). In the absence of CAR-macrophage, this model is described as the one-dimensional ordinary differential equation 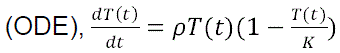 (Model 1). *T*(*t*) represents Raji cell concentration with respect to time *t*. Parameter *p* represents per capita growth rate; *K* controls natural saturation of tumour cell replication at high tumour cell concentration.

To establish a quantitative relationship between Raji cell concentration and CAR-macrophage/ normal macrophage concentration, we developed four models for phagocytosing efficiency: (a) unsaturated phagocytosing efficiency 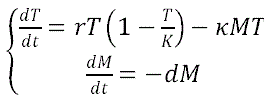 (Model 2), (b) semi-saturated phagocytosing efficiency as Raji cell count increase 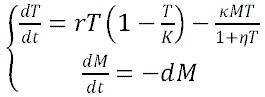^1+l]-^ (Model 3), (c) semi-saturated phagocytosing efficiency as CAR-macrophage/ normal macrophage count increase 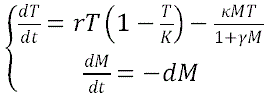 (Model 4), and (d) saturated phagocytosing efficiency 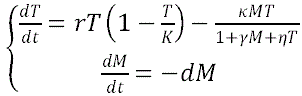 (Model 5). In these models, parameter *Η* represents phagocytosing rate of CAR macrophage/ normal macrophage. *Η* controls the saturation in phagocytosing efficiency as tumour cell concentration increases; *γ* controls the saturation in phagocytosing efficiency as CAR-macrophage/ normal macrophage concentration increases.

To investigate whether phagocytosing Raji cells leads to additional CAR-macrophage death, the change of rate of Raji cells and CAR-macrophages are 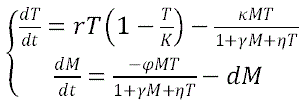, (Model 6). Parameter *φ* represents additional CAR-macrophage cell death rate due to phagocytosing Raji cells.

### Parameter estimation

For the estimation of Raji cell replication parameters, the experimental Raji cell concentration was determined by the formula: Raji cell=TG×10^5^ /ml, where TG represented the number of tumour cells presented in Fig. 4 of Reference [39]. The sum of squared error (SSE) was defined as 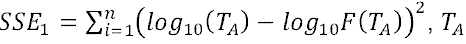 and *F*(*T_A_*) represented experimental Raji cell concentration at time (*t*) and estimated Raji cell concentration at time (*t*). Initial guesses used for parameter estimation were *p*_0_ = 10^-1^ and *K*_0_ = 10^5^.

For the estimation of phagocytosis parameters, the experimental Raji cell concentration was calculated using the formula: Raji cell=2 × TN/ml, where TN represented the number of Raji cell in each well. The CAR-macrophage or normal macrophage concentration was calculated using the formula: macrophage cell=2 × MN/ml, where MN represented the number of Raji cell in each well. The sum of squared error (SSE) was defined as 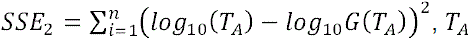 and *G*(*T_A_*) represented experiment Raji cell concentration at time (*t*) and estimated Raji cell concentration at time (*t*). Initial guesses used for parameter estimation were κ_0_ = 10^-1^, y_0_ = and *η*_0_ = 10^-1^.

Next, the Akaike Information Criterion 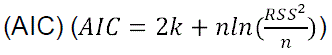 and the modified AIC 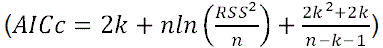 for small sample sizes of unsaturated and saturated neutralisation models were calculated, where *κ* was the number of parameters, *η* was the sample size, and RSS represented the residual sum of squares.

### FACS-based phagocytosis assay

The phagocytosis assay data was sourced from Reference [17]. Briefly, a total of of the complete medium consisting of 8 × 10^5^, 4× 10^5^, 2× 10^5^ transduced CAR-macrophage or normal macrophage were added to 24-well tissue culture plates with 2 × 10^5^ Raji cells and incubated for 24 hours and 48 hours at 35°C in 5% CO_2_. After 24 and 48 hours coculture, the remaining number of Raji cells was analysed by Fluorescence-Activated Cell Sorting (FACS) [17].

## Data Availability

https://github.com/ShlomoBergg/CAR-macrophage-dosage-and-Raji-19

## Acknowledgements

M.L. was supported by Natural Science Foundation of China (82204265), the Science and Technology Innovation Fund of Shenzhen (JCYJ20210324101400001), Key R&D Program of Hunan Province (2023SK2082). S.X. was supported by a fellowship from Lady Davis Fellowship Trust at Technion-Israel Institute of Technology.

## Notes

### Competing Interest Statement

The authors have declared no competing interest.

